# Magnitude and factors associated with missed Human Papillomavirus vaccination opportunities among adolescents in Dar es Salaam, Tanzania

**DOI:** 10.64898/2026.05.15.26353360

**Authors:** Mwajuma Mtandika, Frederick J Kilindo, Fundi Fransiscko, Anthony Kapesa, Basinda Namanya, Dismas Matovelo

## Abstract

**Background:** Tanzania introduced the human papillomavirus (HPV) vaccine in 2018 for girls aged 9–14 years; however, coverage remains suboptimal. Missed opportunities (MOs) for vaccination are an important but understudied barrier, particularly in urban settings. This study assessed factors associated with MOs and explored healthcare providers’ perspectives on barriers and potential solutions in Dar es Salaam.

**Methods:** An embedded mixed-methods study was conducted in public health facilities in Temeke Municipal Council from June - July 2025. The quantitative component involved a cross-sectional survey of 252 parents or caregivers of eligible adolescent girls using structured exit interviews. The qualitative component included in-depth interviews with 20 healthcare providers using a phenomenological approach. Multivariable logistic regression identified factors associated with MOs. Qualitative data were analyzed thematically using Braun and Clarke’s framework.

**Results:** The prevalence of MOs for HPV vaccination was 71.4%. Factors independently associated with MOs included caregiver age ≥40 years (aOR 1.87, 95% CI: 1.02–3.42), female caregiver gender (aOR 1.61, 95% CI: 1.00–2.59), primary education (aOR 2.14, 95% CI: 1.03– 4.45), married status (aOR 1.72, 95% CI: 1.01–2.94), and receiving care at health centers or dispensaries versus hospitals (aOR 1.83, 95% CI: 1.05–3.19). Qualitative findings identified key drivers of MOs, including limited caregiver knowledge, vaccine hesitancy, time constraints, failure to routinely offer vaccination, stock-outs, poor documentation, high workload, and limited outreach. Proposed strategies included routine eligibility screening, reminder systems, community engagement, and supportive supervision.

**Conclusion:** MOs for HPV vaccination are highly prevalent and driven by both caregiver and health system factors. Strengthening routine screening, reminder systems, community engagement, and supervision may improve vaccine uptake.

## INTRODUCTION

Human papillomavirus (HPV) is the most common sexually transmitted infection worldwide and a well-established etiological agent for cervical, anal, and oropharyngeal cancers (1). Cervical cancer remains a major public health problem in low- and middle-income countries (LMICs), accounting for a disproportionate share of global morbidity and mortality. In Tanzania, it is the leading cause of cancer-related deaths among women, with over 10,000 new cases and approximately 7,000 deaths reported annually (2). Prophylactic HPV vaccination is highly effective, preventing more than 90% of HPV-related cancers and precancerous lesions, and constitutes a cornerstone of cervical cancer prevention strategies (3).

Tanzania introduced the HPV vaccine into its national immunization program in April 2018, targeting girls aged 9–14 years through routine facility-based services and outreach platforms, including schools and mobile clinics (4). Initially implemented as a two-dose schedule, the program encountered substantial attrition between doses, largely due to school dropout, school transitions, and limited follow-up mechanisms, resulting in significant missed opportunities (MOs) for complete immunization (5, 6). In response to accumulating evidence and updated WHO recommendations, Tanzania transitioned to a single-dose HPV schedule, a policy shift intended to simplify delivery and improve population-level coverage (7).

This transition aligns with the WHO global strategy for cervical cancer elimination, which calls for achieving 90% HPV vaccination coverage among girls by age 15 by 2030 (8). Although Tanzania reported national first-dose coverage of 98% in 2024, marked subnational disparities persist. Coverage in the Kilimanjaro Region reached 93%, whereas Dar es Salaam reported only 55%, with Temeke Municipality documenting particularly low coverage at 35.2% (9, 10). These gaps have been attributed to a combination of limited caregiver awareness, vaccine hesitancy, health system inefficiencies, logistical constraints, and inconsistent vaccine availability (11, 12). The WHO defines a missed opportunity for vaccination as any health service contact during which an eligible individual does not receive a vaccine dose for which they are eligible, in the absence of contraindications (13). For HPV vaccination, missed opportunities may occur during curative visits, preventive consultations, or routine facility contacts when adolescents accompany caregivers. Contributing factors include providers’ failure to assess vaccination eligibility, vaccine stockouts, rigid service schedules, inadequate documentation, and caregiver resistance or misinformation (14).

Despite the relevance of missed opportunities to HPV vaccination performance, evidence from Tanzania remains limited. Existing studies have largely focused on vaccine uptake or acceptability and often exclude adolescents presenting for non-immunization services or apply narrow operational definitions of MOs (10, 15, 16). To address these gaps, this study assessed the magnitude and determinants of missed opportunities for HPV vaccination among adolescent girls aged 9–14 years attending public health facilities in Temeke Municipal Council, Dar es Salaam. It explored healthcare providers’ perspectives on strategies to reduce missed opportunities and improve vaccine coverage.

## METHODS

### Study Design, Population, and Settings

An embedded mixed-methods design was used in this study, conducted in public health facilities in the Temeke Municipal Council, Dar es Salaam from 1^st^ June to 1^st^ July 2025. This hospital-based cross-sectional study employed a mixed-methods approach to assess missed opportunities (MOs) for HPV vaccination among adolescent girls aged 9–14 years. Quantitative data were collected through client exit interviews with 252 parents or caregivers attending selected public health facilities. Parents or caregivers accompanying adolescent girls aged 9-14 years who received any health service from the selected health facility were selected consecutively until the sample size was met. The phenomenology approach. Qualitative data were obtained through in-depth interviews (IDs) with 20 healthcare providers involved in HPV vaccination service delivery, selected using convenience sampling until saturation was reached. The mixed-methods design enabled triangulation of findings and provided contextual understanding of missed opportunities, consistent with WHO-recommended approaches for evaluating vaccination gaps. The study was conducted in Temeke Municipal Council, an urban district in Dar es Salaam with an estimated population exceeding 1.6 million and approximately 150 health facilities (https://www.temekemc.go.tz/storage/app/uploads/public/59d/ba8/62e/59dba862ed84d383294414.pdf). Data collection was conducted in 40 public health facilities, comprising 3 hospitals, 10 health centers, and 27 dispensaries. Temeke was purposively selected for its consistently low HPV vaccination coverage and its central role in Tanzania’s national HPV vaccine rollout(10, 17).

### Sample Size and Sampling Techniques

For the quantitative component, the sample size was calculated using a single-population proportion formula, assuming a 95% confidence level, a 5% margin of error, and an estimated prevalence of missed opportunities for HPV vaccination of 18% (18). This yielded a minimum sample of 227 participants. After adjusting for a 10% non-response rate, the final sample size was 252 parents or caregivers of adolescent girls aged 9–14 years. For the qualitative component, sample size was guided by thematic saturation. Twenty healthcare providers, including nurses, clinicians, medical attendants, and facility in-charges, were purposively selected based on their involvement in reproductive and child health (RCH) services and HPV vaccination activities.

Simple random sampling was used to select caregivers for exit interviews, while purposive sampling ensured the inclusion of key informants with relevant experience in service delivery.

### Data Collection and Analysis

Quantitative data were collected using a structured, pre-tested questionnaire adapted from the WHO Behavioural and Social Drivers of Vaccination (BeSD) framework (19) and prior empirical studies. The questionnaire was developed in English, translated into Kiswahili, and back-translated to ensure linguistic and conceptual equivalence. It was pilot tested in one facility outside the study area to refine clarity and reliability. The tool captured socio-demographic characteristics, HPV awareness, vaccine offer and uptake, and client- and provider-related factors contributing to missed opportunities. Face-to-face interviews with caregivers were conducted at RCH clinics.

Qualitative data were collected using interview guides. Interviews were conducted in Swahili, audio-recorded with participant consent, transcribed verbatim, and translated into English using a forward–backward translation process. Thematic analysis followed Braun and Clarke’s six-step framework (20) and was supported by NVivo version 14. Quantitative data were entered, cleaned, and analyzed using SPSS version 25. Descriptive statistics summarized participant characteristics and vaccination processes, while multivariate logistic regression identified factors independently associated with missed opportunities. Adjusted odds ratios (AORs) with 95% confidence intervals were reported, and statistical significance was set at p < 0.05.

### Ethics Considerations

Ethical approval was obtained from the CUHAS/BMC Research Ethics Committee (CREC: 952/2025). Administrative clearance was granted by the Dar es Salaam Regional Administrative Secretary (Ref: EA.260/307/028/351). Written informed consent was obtained from all participants before data collection. Participation was voluntary, confidentiality and anonymity were assured, and no personal identifiers were recorded. All procedures adhered to national research regulations and international ethical guidelines for research involving human participants.

## RESULTS

### A: Quantitative Results

#### Demographic Characteristics of Parents/Caregivers

A total of 252 parents and caregivers of adolescent girls aged 9–14 years participated in the study across 40 public health facilities in the Temeke Municipal Council, Dar es Salaam. The majority were recruited from health centres (n=134, 53.2%), followed by dispensaries (n=68, 27.0%), district hospitals (n=32, 12.7%), and regional referral hospitals (n=18, 7.1%). Over two-thirds of the participants were female (n= 184, 73.0%), and the sample was predominantly Muslim (n= 176, 69.8%). Most respondents had completed only primary education (n= 208, 82.5%) and were residents of the Temeke Municipal Council. Petty trading was the most common occupation, reported by 216 participants (85.7%). The majority of respondents were mothers of the adolescent girls (n= 192, 76.2%), followed by fathers (n= 36, 14.3%) and other relatives (n= 24, 9.5%) (Table 1).

**Table 1:** Demographic Characteristics of Parents/Caregivers (N= 252)

| Variable | Category | Frequency<br>(n) | Percent (%) |
| --- | --- | --- | --- |
| **Health Facility Type** | Regional Referral Hospital | 18 | 7.1 |
|  | District Hospital | 32 | 12.7 |
|  | Health Centre | 134 | 53.2 |
|  | Dispensary | 68 | 27.0 |
| **Gender** | Male | 68 | 27.0 |
|  | Female | 184 | 73.0 |
| **Age (years)** | Mean ± SD | — | 39.7 ± 7.2 |
| **Religion** | Christian | 70 | 27.8 |
|  | Muslim | 176 | 69.8 |
|  | Others | 6 | 2.4 |
| **Marital Status** | Single | 58 | 23.0 |
|  | Married | 158 | 62.7 |
|  | Widowed/Divorced | 36 | 14.3 |
| **Education Level** | No Formal Education | 12 | 4.8 |
|  | Primary School | 208 | 82.5 |
|  | Secondary School | 24 | 9.5 |
|  | Diploma/University | 8 | 3.2 |
| **Occupation** | Petty Trader | 216 | 85.7 |
|  | Formal Employment | 18 | 7.1 |
|  | Casual Labor | 10 | 4.0 |
|  | Others* | 8 | 3.2 |
| **Relationship to Child** | Mother | 192 | 76.2 |
|  | Father | 36 | 14.3 |
|  | Other Relative | 24 | 9.5 |
| **Adolescent Age (years)** | Median (IQR) | — | 12 (10–13) |
| **Adolescent Education** | Primary School | 226 | 89.7 |
|  | Secondary School | 18 | 7.1 |
|  | No Formal Education | 8 | 3.2 |
| **Reason for Visit** | Medical Consultation (sick) | 132 | 52.4 |
|  | Health Check-up | 56 | 22.2 |
|  | Vaccination | 42 | 16.7 |
|  | Accompanying Only | 14 | 5.6 |
|  | Hospitalization | 8 | 3.2 |

\* “Others” under occupation included small-scale farmers, tailors, food vendors, and domestic workers as reported by participants during interviews.

#### Magnitude of Missed Opportunities for HPV Vaccination

Among the 252 caregivers interviewed, 180 adolescent girls (71.4%) experienced at least one missed opportunity to receive HPV vaccination during their health facility visit. Although 58.7% of caregivers received some information about HPV vaccination and 52.4% were asked about their child’s vaccination status, fewer than half were offered the vaccine. Only 28.6% of eligible girls were vaccinated during the visit, reflecting substantial gaps in service integration and provider engagement.

Additionally, only 68 (27.0%) caregivers were informed of the vaccine administered, 56 (22.2%) were told the date of the next appointment, and 44 (17.5%) received a written record of the vaccination. While 84 (33.3%) were given information about possible side effects, only 56 (22.2%) correctly recalled them. Overall, based on the study’s definition of a missed opportunity as any instance in which an eligible adolescent girl did not receive the HPV vaccine despite access to services, 180 (71.4%) of adolescent girls experienced at least one missed opportunity during the visit (Table 2).

**Table 2:** Magnitude of Missed Opportunities for HPV Vaccination (N= 252)

| **Variables** | **Yes, N (%)** | **No, N (%)** |
| --- | --- | --- |
| The provider shared HPV information | 148 (58.7) | 104 (41.3) |
| The provider asked HPV vaccination status | 132 (52.4) | 120 (47.6) |
| Offered the HPV vaccine | 112 (44.4) | 140 (55.6) |
| Accepted vaccine (if offered) | 98 (87.5) | 14 (12.5) |
| Child vaccinated today | 72 (28.6) | 180 (71.4) |
| Told which vaccine was given | 68 (27.0) | 184 (73.0) |
| Told the date of the next appointment | 56 (22.2) | 196 (77.8) |
| Appointment recorded in writing | 44 (17.5) | 208 (82.5) |
| Received information on side effects | 84 (33.3) | 168 (66.7) |
| Correctly mentioned side effects | 56 (22.2) | 196 (77.8) |

#### Reported factors contributing to missed Opportunities for HPV Vaccination

Among the 180 caregivers whose daughters were not vaccinated during the visit, both client- and provider-related factors contributed to missed opportunities. The leading client-related factor was limited knowledge about HPV or the vaccine, reported by 98 (54.4%) caregivers. This was followed by not visiting the facility specifically for vaccination (84, 46.7%), insufficient time to wait for services (62, 34.4%), and religious or personal objections to vaccination (38, 21.1%). Additional reasons included challenges in accessing the vaccination site (22, 12.2%), forgetting the appointment (18, 10.0%), concerns about previous vaccine reactions (14, 7.8%), mistrust of health workers or vaccines (10, 5.6%), and community discouragement (6, 3.3%). On the provider side, the most frequently reported barrier was that health workers did not mention HPV vaccination during the visit (108, 60.0%). Other challenges included stock-outs of vaccines 52 (28.9%), being informed that the child was not eligible 26 (14.4%), lack of syringes or other vaccination supplies 18 (10.0%), closed vaccination areas 14 (7.8%), absence of the vaccinator 10 (5.6%), long waiting times 8 (4.4%), and poor treatment by staff 6 (3.3%) (Table 3).

**Table 3:** Factors Contributing to Missed Opportunities for HPV Vaccination (N= 180, multiple responses allowed)

| **Client-Related Factors** | **Frequency (N)** | **Percentage (%)** |
| --- | --- | --- |
| Lack of knowledge about HPV or the vaccine | 98 | 54.4 |
| Did not come for vaccination | 84 | 46.7 |
| No time to wait | 62 | 34.4 |
| Religious/personal beliefs against vaccination | 38 | 21.1 |
| Difficult to access the vaccination site | 22 | 12.2 |
| Forgot appointment | 18 | 10.0 |
| Previous vaccine reaction | 14 | 7.8 |
| Mistrust of health workers or vaccines | 10 | 5.6 |
| The community discourages vaccination | 6 | 3.3 |
| **Provider-Related Factors** | **Frequency (N)** | **Percentage (%)** |
| The health worker did not mention HPV vaccination | 108 | 60.0 |
| No vaccines available | 52 | 28.9 |
| Told the child not eligible | 26 | 14.4 |
| No syringes or supplies | 18 | 10.0 |
| The vaccination area was closed | 14 | 7.8 |
| Vaccinator was absent | 10 | 5.6 |
| Long waiting time | 8 | 4.4 |
| Poor treatment by the provider | 6 | 3.3 |
| Referred to another facility for vaccination | 42 | 23.3 |

**Table 4:** Multivariate Logistic Regression Results for Factors Associated with Missed Opportunities for HPV Vaccination (N= 252)

| Variable | MO Present<br>(n, %) | MO Absent<br>(n, %) | Unadjusted<br>OR (95% CI) | p-value | Adjusted<br>OR (95% CI) | p-value |
| --- | --- | --- | --- | --- | --- | --- |
| **Age Group** |  |  |  |  |  |  |
| <35 years | 78 (65.5) | 41 (34.5) | 1.00 |  | 1.00 |  |
| ≥40 years | 102 (76.7) | 31 (23.3) | 1.94 (1.12–3.36) | 0.017 | 1.87 (1.01–3.45) | 0.042 |
| **Gender** |  |  |  |  |  |  |
| Male | 42 (61.8) | 26 (38.2) | 1.00 |  | 1.00 |  |
| Female | 138 (75.0) | 46 (25.0) | 1.68 (1.01–2.80) | 0.045 | 1.61 (1.00–2.60) | 0.049 |
| **Marital Status** |  |  |  |  |  |  |
| Single | 34 (58.6) | 24 (41.4) | 1.00 |  | 1.00 |  |
| Married | 118 (74.7) | 40 (25.3) | 1.76 (1.03–3.01) | 0.038 | 1.72 (1.01–2.94) | 0.046 |
| **Education Level** |  |  |  |  |  |  |
| Secondary or higher | 12 (42.9) | 16 (57.1) | 1.00 |  | 1.00 |  |
| Primary | 158 (76.0) | 50 (24.0) | 2.21 (1.08–4.52) | 0.030 | 2.14 (1.02–4.47) | 0.041 |
| **Occupation** |  |  |  |  |  |  |
| Formal employment | 10 (55.6) | 8 (44.4) | 1.00 |  | 1.00 |  |
| Petty trader | 158 (73.1) | 58 (26.9) | 2.05(1.01–4.17) | 0.057 | 1.98 (1.00–3.91) | 0.06 |
| **Facility Type** |  |  |  |  |  |  |
| Hospital | 28 (58.3) | 20 (41.7) | 1.00 |  | 1.00 |  |
| Health Centre/<br>Dispensary | 106 (79.1) | 28 (20.9) | 1.89(1.10–3.25) | 0.021 | 1.83 (1.05–3.19) | 0.034 |

#### Factors Associated with Missed Opportunities for HPV Vaccination

Bivariate analysis revealed several caregiver characteristics significantly associated with missed opportunities (MOs) for HPV vaccination. Caregivers aged ≥40 years were more likely to report MOs than those aged < 35 years (OR= 1.94, 95% CI: 1.12–3.36, p= 0.017). Female caregivers had higher odds of MOs than males (OR= 1.68, 95% CI: 1.01–2.80, p= 0.045), and those with primary education were significantly more likely to experience MOs than those with secondary or higher education (OR= 2.21, 95% CI: 1.08–4.52, p= 0.030). Marital status was also significantly associated, with married caregivers more likely to report MOs than single caregivers (OR= 1.76, 95% CI: 1.03–3.01, p= 0.038). Facility type was a notable predictor, with caregivers attending health centres/dispensary levels having higher odds of MOs than those visiting hospitals (OR= 1.89, 95% CI: 1.10–3.25, p= 0.021).

Multivariate logistic regression confirmed the significance of several predictors. Age ≥40 years (aOR= 1.87, 95% CI: 1.01–3.45, p= 0.042), female gender (aOR= 1.61, 95% CI: 1.00–2.60, p= 0.049), primary education level (aOR= 2.14, 95% CI: 1.02–4.47, p= 0.041), and being married (aOR= 1.72, 95% CI: 1.01–2.94, p= 0.046) remained statistically significant. Facility type was also a strong predictor, with caregivers attending health centers/dispensaries more likely to report MOs (aOR= 1.83, 95% CI: 1.05–3.19, p= 0.034). Religion and occupation showed no statistically significant association with missed opportunities in either bivariate or multivariate analysis.

### B. Qualitative Findings

The qualitative findings from this study revealed critical insights into the systemic, behavioral, and contextual factors contributing to missed opportunities for HPV vaccination among adolescent girls in the Temeke Municipal Council.

#### Demographic Characteristics of Healthcare Workers (HCWs) Interviewed (N= 20)

| Variable | Category | Frequency (N) |
| --- | --- | --- |
| Sex | Male | 8 |
|  | Female | 12 |
| Age (years) | Mean ± SD | 39.8 ± 7.4 |
| Distribution of HCWs | Hospital | 6 |
|  | Health Centre | 8 |
|  | Dispensary | 6 |
| Marital Status | Married | 14 |
|  | Single | 4 |
|  | Divorced/Widowed | 2 |
| Education Level | Diploma | 10 |
|  | University Degree | 8 |
|  | Secondary/Advanced School | 2 |
| Cadre | Nurse | 12 |
|  | Clinical Officer | 5 |
|  | Medical Doctor | 3 |
| Department/Section | Reproductive & Child Health (RCH) | 9 |
|  | Outpatient | 5 |
|  | Family Planning | 3 |
|  | ANC/Emergency/Other | 3 |
| Years of Experience | <5 years | 6 |
|  | 5–10 years | 8 |
|  | >10 years | 6 |
| Trained on VPDs | Yes | 14 |
|  | No | 6 |

Through in-depth interviews with healthcare providers across hospitals, health centres, and dispensaries, four major themes emerged: provider knowledge and communication; service integration challenges; barriers to vaccine uptake; and proposed strategies for improvement.

Healthcare providers demonstrated basic awareness of HPV and its link to cervical cancer, but knowledge gaps persisted. While most understood the target age group, few could confidently explain the rationale for early vaccination. As one clinical officer noted, *“We vaccinate girls aged 9–14, but some staff still ask why so young*.*”* A medical doctor added, *“I know HPV is linked to cervical cancer, but not everyone here can explain it clearly to parents*.*”* Providers acknowledged their role in educating caregivers but felt constrained by time and workload, with a nurse stating, *“We’re supposed to inform every parent, but the workload makes it hard*.*”*

Service delivery challenges were a recurring theme, particularly around poor integration of HPV vaccination into routine care. Many providers admitted that eligible girls were not consistently screened during non-immunization visits. A health assistant explained, *“Girls come for other services, but unless it’s an immunization day, we don’t check their vaccination status*.” Stock-outs and documentation gaps further disrupted continuity, as one nurse shared, *“We run out of HPV vaccines often, especially during school holidays,”* while a clinical officer added, “We don’t have a system to track who missed the vaccine.”

Barriers to vaccine uptake were identified at both client and provider levels. Caregivers often lacked awareness or expressed hesitancy, influenced by cultural beliefs and misinformation. A nurse observed, *“Some parents say they’ll come back, but they never do*,” while a health assistant noted, “*Unless the parent asks, we don’t always mention HPV*.” Providers also cited rigid clinic schedules and assumptions about eligibility as contributors to missed opportunities. As one medical doctor put it, “*HPV is rarely discussed unless it’s scheduled, and some staff assume the child isn’t eligible*.*”*

Despite these challenges, providers proposed actionable strategies to reduce MOs. Routine screening, use of triage tools, and reminder systems were widely recommended. A clinical officer emphasized, *“We need to ask about HPV status for every adolescent visit*,” while a nurse suggested, *“Parents forget. A simple reminder would help*.*”* Community engagement was also seen as vital, with a medical doctor stating, *“When teachers talk about HPV, parents listen*,” and a facility in charge adding, *“We need refresher training—guidelines change, and we’re left behind*.*”* These insights underscore the need for integrated, provider-supported, and community-driven approaches to improve HPV vaccine uptake.

## DISCUSSION

This study demonstrates a high burden of missed opportunities for HPV vaccination in the Temeke Municipal Council, with more than seven in ten eligible adolescent girls failing to receive the vaccine despite accessing health services. The observed magnitude is concerning and underscores persistent weaknesses in opportunistic vaccination delivery in urban health facilities. Quantitative findings revealed that fewer than half of caregivers were offered HPV vaccination, while qualitative data highlighted poor integration of HPV services into routine care, limited screening during non-immunization visits, and fragmented documentation practices.

Comparable patterns have been reported in Sub-Saharan Africa (SSA) and other Tanzanian settings, where inadequate service integration and limited provider engagement contributed to missed opportunities exceeding 60% (10, 11, 21). In Kilimanjaro’s Hai District, uptake was estimated at 65%, constrained by misinformation and lack of sensitization (12), while the HPV-Plus program demonstrated that comprehensive health education could increase acceptance despite persistent hesitancy linked to inadequate caregiver involvement (18, 21). These parallels indicate that missed opportunities in Temeke reflect broader systemic challenges across urban Tanzania, underscoring the importance of improving service integration and provider engagement. In contrast, lower rates reported in Rwanda and Ethiopia (25–35%) have been attributed to strong school-based delivery models, integrated screening systems, and structured digital tracking tools (14, 22, 23). Conversely, in more resource-constrained contexts, such as Nigeria and Malawi, MO rates exceed 80%, largely due to stock-outs, weak infrastructure, and entrenched sociocultural resistance (24, 25). These comparisons suggest that Temeke’s high MO burden reflects modifiable system-level gaps rather than entrenched caregiver resistance.

Caregiver knowledge and perceptions emerged as important determinants of missed opportunities. Limited awareness and misconceptions, particularly the belief that HPV vaccination is only relevant for sexually active girls, were more common among caregivers with lower education levels. Similar findings have been documented in SSA at large and in other parts of Tanzania, such as in Kilimanjaro and in Ilala, Dar es Salaam (10, 11, 16). However, evidence from out-of-school and in-school-based programs in SSA suggests that delivery platforms that reduce reliance on caregiver initiative can mitigate the influence of HPV vaccination uptake (26).

On the supply side, provider failure to mention HPV vaccination during visits was the most frequently reported barrier, affecting 60% of missed cases. Stock-outs, uncertainty about eligibility, and weak referral mechanisms further compounded missed opportunities. These findings mirror challenges reported in Malawi and Nigeria (24, 25), but contrast with experiences from Ethiopia, where structured supervision and integrated workflows have minimized provider-related lapses (22).

Importantly, healthcare providers in Temeke articulated clear, feasible strategies to reduce missed opportunities, including routine eligibility screening at all service points, reminder systems, strengthened documentation, and enhanced community engagement. However, implementation was constrained by high workloads, limited staffing, inconsistent supervision, and supply chain interruptions. Addressing these operational barriers is critical to translating provider motivation into sustained improvements in vaccine delivery. Weak referral and follow-up systems further exacerbated missed opportunities, as few unvaccinated girls were directed to alternative service points. Taken together, these findings underscore that missed opportunities arise from a complex interplay between caregiver knowledge and motivation, provider practices, and systemic limitations.

These gaps reflect patterns similar to those seen in Kilimanjaro and elsewhere in SSA, where the lack of integrated screening contributed to frequent MOs (16, 26). In contrast, several SSA countries have embedded screening into routine workflows, thereby reducing reliance on provider memory (22, 23). Reminder tools such as appointment cards, short messaging services (SMS), and verbal prompts were also recommended and shown to be effective when applied consistently, and reminder systems improved vaccine completion (22, 23). However, concerns about unreliable mobile communication in low-income settings resonate with experiences in Sierra Leone (27), suggesting that reminder systems must be complemented with community-based follow-up. Community engagement through schools, community health workers, and religious leaders was highlighted as a powerful facilitator, consistent with evidence from Tanzania’s HPV-Plus program (21) and Rwanda’s sustained school and faith-based collaborations (26). Training and supportive supervision were also prioritized by providers, who emphasized the need for regular refresher courses on HPV guidelines and communication. However, inconsistent and often punitive supervisory practices in Temeke limited effectiveness, which may be improved by collaborative supervision. Overall, while providers expressed a strong commitment, operational challenges, such as high client volumes, limited staffing, stock-outs, and weak supervisory structures, constrained the systematic implementation of these strategies.

In conclusion, this study demonstrates that missed opportunities for HPV vaccination in Temeke Municipal Council are widespread and shaped by both demand- and supply-side factors, echoing challenges observed in other Tanzanian districts while also reflecting unique contextual dynamics. Compared with regional peers, Temeke’s rates are higher than those of Rwanda and Ethiopia (22, 26), where stronger system integration has minimized MOs, but lower than in some resource-constrained settings such as Nigeria, Malawi, and Sierra Leone (24, 25, 27). Importantly, caregiver acceptance and provider motivation remain relatively strong, suggesting that system-level interventions, particularly integration of routine screening, structured reminders, consistent supervision, and community engagement, could significantly improve uptake.

The strengths and limitations of this study merit mention. The mixed-methods design is a key strength of this study, enabling quantification of missed opportunities while providing contextual insights into underlying mechanisms. The inclusion of diverse facility types enhanced representativeness in the urban setting. However, the cross-sectional design limits causal inference, and reliance on self-reported data may have introduced recall and social desirability bias. The study’s focus on a single urban municipality limits generalizability to rural settings, and the exclusion of adolescent and community leader perspectives constrained deeper system-level analysis.

However, the findings highlight the need for national strategies that institutionalize provider training, improve logistics, and embed HPV vaccination into broader adolescent health packages, while simultaneously expanding school- and community-based outreach to counter misconceptions and reduce reliance on caregiver initiative. From a policy perspective, these results call for scaling up integrated digital tracking and reminder tools, strengthening supply chain management, and fostering continuous provider mentorship. For practice, they underscore the importance of empowering providers with tools and supportive supervision to integrate HPV vaccination into all service encounters. For research purposes, further evaluation of hybrid models that combine school-based delivery with opportunistic facility-based vaccination will be critical to optimizing coverage and sustainability. By addressing both structural and behavioral barriers, Tanzania can move closer to equitable HPV vaccination coverage and the reduction of cervical cancer risk among adolescent girls.

## Conclusion

Missed opportunities for HPV vaccination among adolescent girls in Temeke Municipal Council are highly prevalent and driven by intersecting caregiver, provider, and system-level factors. Despite these challenges, caregiver acceptance and provider commitment remain relatively strong, indicating substantial potential for improvement through targeted system strengthening. Integrating HPV vaccination into routine service workflows, institutionalizing age-based screening protocols, improving vaccine logistics, and implementing reminder systems are essential to reducing missed opportunities.

## Data Availability

Data from this study (S1 Appendix) are attached in the support information section of this article.

## Acknowledgment

Special thanks to all caretakers and healthcare providers in the Temeke Municipal Council who voluntarily participated in the study.

## Author’s contributions

MM, DM, and BN: conceived, designed, and executed the study, MM and FJK: participated in data and sample collection, MM, DM and FF: involved in data analysis and interpretation, DM and BN: supervised the execution of the study, MM and DM: prepared the first manuscript which was critically reviewed and approved by all authors.

## Abbreviations

HPV: Human Papillomavirus
LMICs: Low-Middle-Income Countries
SMS: Short messaging service
MOs: Missed opportunities
PAPM: Precaution Adoption Process Model
SSA: Sub-Saharan Africa
WHO: World Health Organization

## Availability of data and materials

Data from this study (S1 Appendix) are attached in the support information section of this article.

